# Associations of Water Quality with Cholera in Case-Control Studies: A Systematic Review and Meta-Analysis

**DOI:** 10.1101/2023.09.06.23295113

**Authors:** Thuy Tien Nguyen, Chaelin Kim, Gerard Goucher, Jong-Hoon Kim

**Author notes:** Corresponding author Name: Jong-Hoon Kim Postal address: International Vaccine Institute, SNU Research Park, 1 Gwanak-ro, Gwanak-gu, Seoul 08826, South Korea.

## Abstract

Cholera is a significant health risk for low- and middle-income countries (LMIC), and the threat of outbreaks is likely to increase due to climate change. To keep up to date with the link between water quality and cholera, we conducted a systematic review and meta-analysis to update a previous review. We searched Embase, Web of Science and PubMed for literature published between 2016 and 2022. Search terms were consistent with the previous review. Study quality was assessed using Risk Of Bias In Non-randomized Studies - of Exposures (ROBINS-E). Exposures of water were categorized according to the WHO/UNICEF Joint Monitoring Program for Water Supply, Sanitation and Hygiene (JMP) and further divided by the service ladder. Odds ratios were extracted and pooled by performing random-effects meta-analysis. We identified 22 new eligible studies and analysed them together with the 45 studies included in the previous review. Analyses revealed higher odds of cholera when consuming sachet water (OR=1.69, 95% CI: 1.13 to 2.52), unimproved water (OR=2.91, 95% CI: 1.21 to 7.02), surface water (OR=3.40, 95% CI: 2.52 to 4.58), and untreated water (OR=2.51, 95% CI: 2.03 to 3.10). Meanwhile, treating water (OR=0.42, 95% CI: 0.27 to 0.65), by boiling (OR=0.38, 95% CI: 0.17 to 0.84) or chlorination (OR=0.37, 95% CI: 0.17 to 0.83), and drinking basic water (OR=0.44, 95% CI: 0.27 to 0.69) showed protection. Pooled estimates changed with updated evidence while qualitative insights on the protective or risk factors remain valid. Relatively low-cost methods like boiling or chlorinating water provide good protection comparable to providing basic water to the public.

Systematic review registration: PROSPERO 2021 CRD42021271881

## Introduction

Cholera is an acute diarrhoeal disease caused by the bacterium *Vibrio cholerae*. The disease is mainly transmitted through the faecal-oral route by consuming contaminated water or food, or through person-to-person contact [1].

Although cholera is not a newly emerging disease, it remains a significant global public health threat, especially for low- and middle-income countries (LMICs) with poor sanitation infrastructure and limited access to clean water [2]. In 2022, more than 29 countries have reported new outbreaks to the World Health Organization (WHO), with Lebanon and Syria not being considered cholera non-endemic countries [2]. Major outbreaks often occur in areas of humanitarian crisis, political instability, and water insecurity [2]. This is illustrated by the 2016 outbreak in Yemen, one of the largest cholera epidemics in recent times as a result of ongoing armed conflict [3].

Accurately estimating the global burden of cholera is difficult. While the World Health Organization (WHO) reported 323,320 cholera cases and 857 deaths in 2020, these figures are likely to be considerably underestimated [4]. Factors such as inadequate surveillance systems, fears of negative economic impacts on trade and tourism, lack of diagnostic tools, and the recent COVID-19 pandemic may have contributed to the underreporting of cholera cases [5]. A modelling study estimated the cholera burden to be between 1.3 and 4.0 million cases annually in cholera-endemic countries [6]. The burden may increase with climate change putting even high-income countries at risk in future [7, 8]. With extreme weather events such as changing rainfall patterns, recurrent flooding, and rising temperatures, the risk of faecal-oral pathogens contaminating the environment is increasing. This threatens water quality and increases the transmission of waterborne diseases [7, 8]. Larger and more deadly cholera outbreaks have already been observed as a consequence [2].

Main risk factors for cholera outbreaks are inadequate water quality, poor sanitation and hygiene, and overcrowding [1]. These risks can be counteracted by implementing water, sanitation, and hygiene (WASH) interventions. WASH interventions are one of the most important factors in limiting and preventing outbreaks [9, 10]. The importance of WASH is being emphasized by the establishment of the WHO/UNICEF Joint Monitoring Program (JMP) for Water, Sanitation and Hygiene in 1990. The JMP monitors the global progress on WASH towards achieving the Sustainable Development Goals [11]. It regularly collects national and regional-level WASH estimates for more than 190 countries. Data are being reported for four different WASH categories of water, sanitation, hygiene, and menstrual health. The former three comprise five subcategories and cover different settings, such as households, schools, and health facilities [12, 13]. Thus, understanding the WASH association with cholera standardized by JMP categories can provide a way to explore the risk associated with cholera in areas where JMP estimates are available.

Although many have already investigated the association between WASH and cholera [14–17], it is important to regularly update knowledge based on new evidence. A better understanding is crucial to guide decision-making and can lead to more cost-effective adaption of intervention programs and thus strengthen the impact of future cholera prevention programs. To our knowledge, there is currently only one systematic review and meta-analysis that explored the association between WASH exposures and cholera in case-control studies. Wolfe et al. published their study in 2018, evaluating studies published between 1990 and 2016 [17]. The authors included water, sanitation, and hygiene and classified WASH exposures according to the JMP standards [17]. However, the JMP-specific service ladders developed for further grading and specification of WASH categories were not used.

While Wolfe *et al.* examined all three categories of WASH which include water, sanitation, as well as hygiene [17], no systematic review has yet focused exclusively on water quality exposures related to cholera. Out of WASH, focusing on the association between water and cholera is compelling as water is a basic human need and cholera spreads mainly via unsafe water [1]. Case-control studies are particularly interesting to focus on when studying this association as our preliminary search revealed that major evidence was generated in this particular study design as well as its consistency being an advantage for meta-analyses.

The aim of this study is to update the existing systematic review and meta-analysis while focusing on the association between water and cholera. This review will incorporate recent evidence and provide updated insights on the association between water quality and cholera in LMICs while implementing a more rigorous assessment of the study quality.

## Methods

### Search Strategy and Eligibility Criteria

The development of the systematic review and meta-analysis adhered to the Preferred Reporting Items for Systematic Reviews and Meta-Analyses (PRISMA) [18]. To systematically identify relevant peer-reviewed articles PubMed, Web of Science, and Embase were chosen as databases. The search terms were kept consistent with the previous search terms utilized in Wolfe *et al.*: (“case control” OR “case-control” AND “cholera”) [17].

Results were restricted to articles published in English between 01 July 2016 and 02 September 2022 since articles published before 01 July 2016 were already included in the Wolfe *et al.* review [17]. All search records were imported to the software Sciwheel, deduplicated, and further screened according to the following predefined inclusion and exclusion criteria: Case-control studies focusing populations in LMICs were eligible to be included, whereas studies located in high-income countries were excluded. Cases were defined as people infected with cholera, while controls were defined as people not infected with cholera. No restrictions were put on age, gender, and socioeconomic status of the population. Furthermore, studies had to analyse associations between WASH and cholera using odds ratios (OR) to be eligible. Detailed information on the Population, Intervention, Comparison, Outcomes and Study Design (PICOS) criteria [19] applied can be found in the study protocol [20].

### WASH Exposure Categories with Focus on Water

The JMP WASH categories were used in order to classify the exposures more precisely by additionally using the JMP service ladder to further differentiate these categories into finer levels [21]. However, as this study focuses on water quality, only the category of water was included for further analysis. In addition to the JMP category water source, water treatment and water management were included to describe water and its handling in more detail and to be consistent with Wolfe *et al.* [17] (Table 1). Exposures deemed eligible were compared to the JMP service ladder for water source. They were subsequently allocated to one of the five subcategories (safely managed, basic, limited, unimproved, surface water) according to its definition given by the JMP [22]. Water treatment and water management were further divided into treated water and untreated water as well as safe water storage and unsafe water storage, respectively.

**Table 1.**
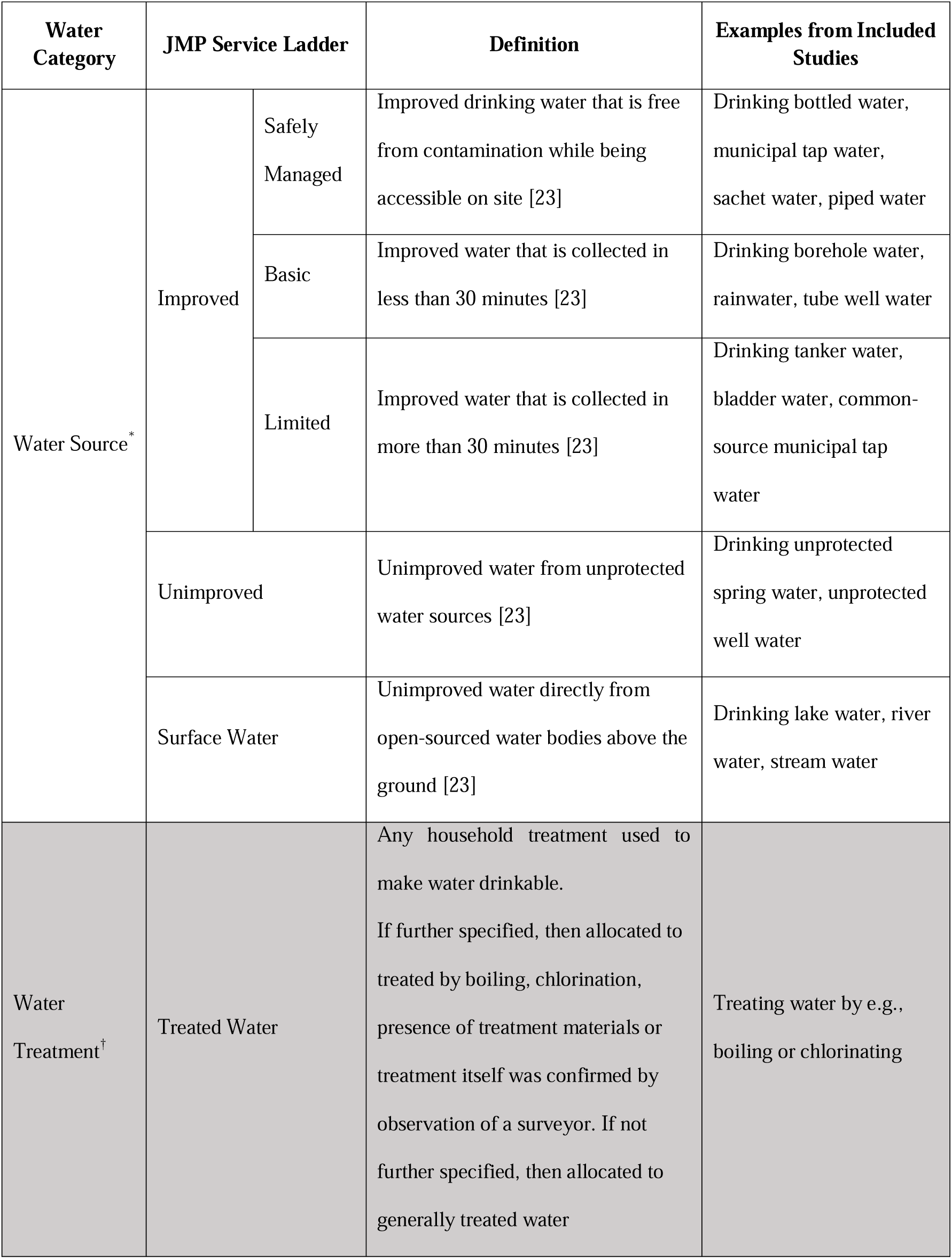
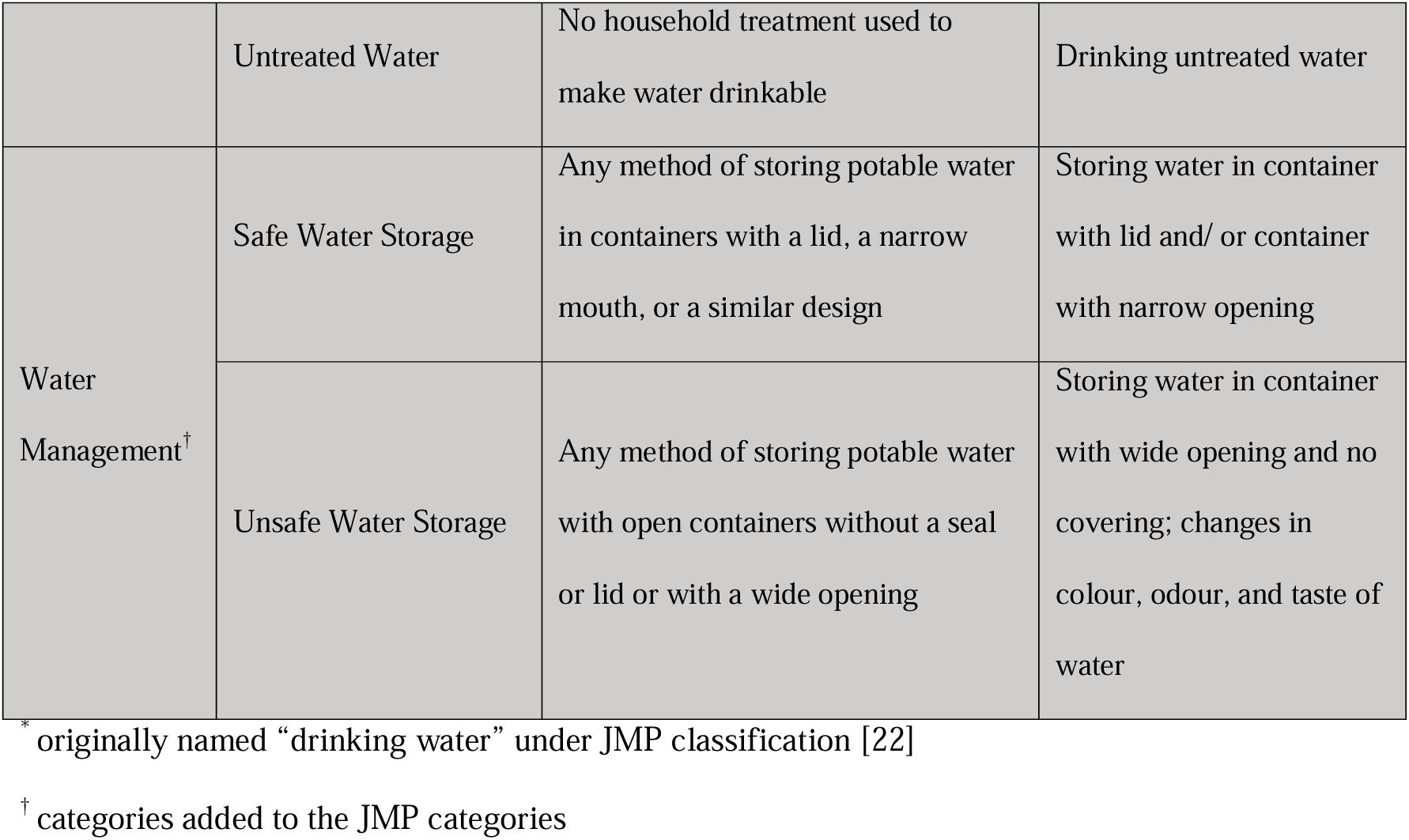
Water related exposures from studies included in the analysis and corresponding JMP service ladder.

Additionally, a more differentiated classification was introduced for similar exposures of the same category. An example would be the differentiation of boiled water and chlorinated water, rather than analysing them jointly as treated water or having to exclude one exposure. Other additional inclusion criteria were developed if this differentiation was not sufficient to avoid unit-of-analysis error and to ensure that certain populations were not overrepresented in the meta-analyses.

### Data Extraction

In addition to the articles selected based on the previous review [17], title and abstract of the newly identified articles were screened and checked for relevance by two independent researchers (CK and TN) and eligibility was determined in accordance with the PICOS framework. After this initial screening, full texts of included studies were further independently examined for eligibility by the same researchers. Discrepancies in both steps were discussed afterwards with a third researcher (J-HK) to reach consensus.

Relevant data were extracted and compiled using Google Sheets. For each article, following information was extracted: the country where the study was conducted, the WASH exposures analysed, the OR and corresponding confidence interval (CI), the general adjustment factors used, and the adjustment for other WASH exposures.

### Risk of Bias Assessment

The quality appraisal of the eligible studies was conducted using the Risk Of Bias In Non-randomized Studies - of Exposures (ROBINS-E) [23]. The tool assesses the risk of bias in seven different domains: 1) confounding, 2) measurement of exposure, 3) selection of participants, 4) post-exposure interventions, 5) missing data, 6) measurement of outcome, and 7) selective reporting of results. Each domain was graded on a scale of “low risk of bias”, “some concerns”, “high risk of bias”, and “very high risk of bias” which was subsequently summarized in an overall risk of bias. Studies scoring “low risk” in more than one domain were categorized as overall “some concerns”; studies scoring “some concerns” in more than five domains were considered “high risk”; and those scoring “high risk” in more than one domain were categorized as “very high risk”.

The assessment of quality was performed by two reviewers independently (CK and TN) and then discussed in conjunction with the third researcher (J-HK).

### Statistical Analysis

Studies labelled as “very high risk” during the quality assessment were excluded from the analysis. Exposures that were identified by the included articles as contaminated and potentially even the origin of an outbreak were excluded. This was due to the high likelihood of bias in the meta-analysis as a result of misclassification of an exposure that was originally confirmed to be contaminated. In addition, only one exposure per study population was included in each analysis to avoid unit-of-analysis error and to ensure that no study population was given more weight in the analysis. Exposures with narrower confidence intervals and thus more precise estimates were chosen over similar exposures with wider intervals. Preference was also given to exposures that were more comparable to other exposures in the same subcategory or subgroup. For example, if an article examined the exposure of “drinking river water” and “drinking swamp water” in the same population, “drinking river water” was included in the analysis and “drinking swamp water” was excluded. Reason being that “drinking river water” was more comparable to the exposures included in other articles in that particular subgroup. Also, confidence intervals had to be reported in the study to be included in the statistical analysis. Exposures with p-values only were excluded.

The statistical programming language R (version 4.2.2) [24] with the *metafor* package (version 3.8.1) [25] was utilized to conduct the meta-analysis. Meta-analysis was performed for categories with at least two studies with the potential of meaningful pooling [26]. Adjusted ORs were preferred over crude ORs; if not given, matched ORs were utilized. If neither the adjusted OR nor the matched OR were reported, the crude OR was used for meta-analysis. Random effects models were chosen to fit the data over the fixed effects model since real differences in the effect sizes, instead of sampling error, were suspected because of the variability in the setting and the population studied. The heterogeneity between studies was estimated by restricted maximum likelihood [27] and calculated using τ^2^ Cochran’s Q*-*statistic for subgroup interactions [28], and I^2^-statistic [29]. An overall summary estimate with confidence interval was calculated for each JMP subcategory and two additional categories: “safely managed water” and “treated water”. Publication bias was primarily inspected by examining symmetry of the funnel plots and calculating Egger’s regression [30].

## Results

### Study Selection

The search yielded a total number of 206 articles consisting of 86 articles from PubMed, 55 from Web Of Science, and 65 from Embase published between 01 July 2016 and 02 September 2022 (Fig 1). After deduplication, 120 unique articles were left for title and abstract review. This subsequently resulted in 36 articles that were deemed eligible for full text screening. During the full text screening, 14 articles were further excluded (Fig 1).

**Fig 1.**
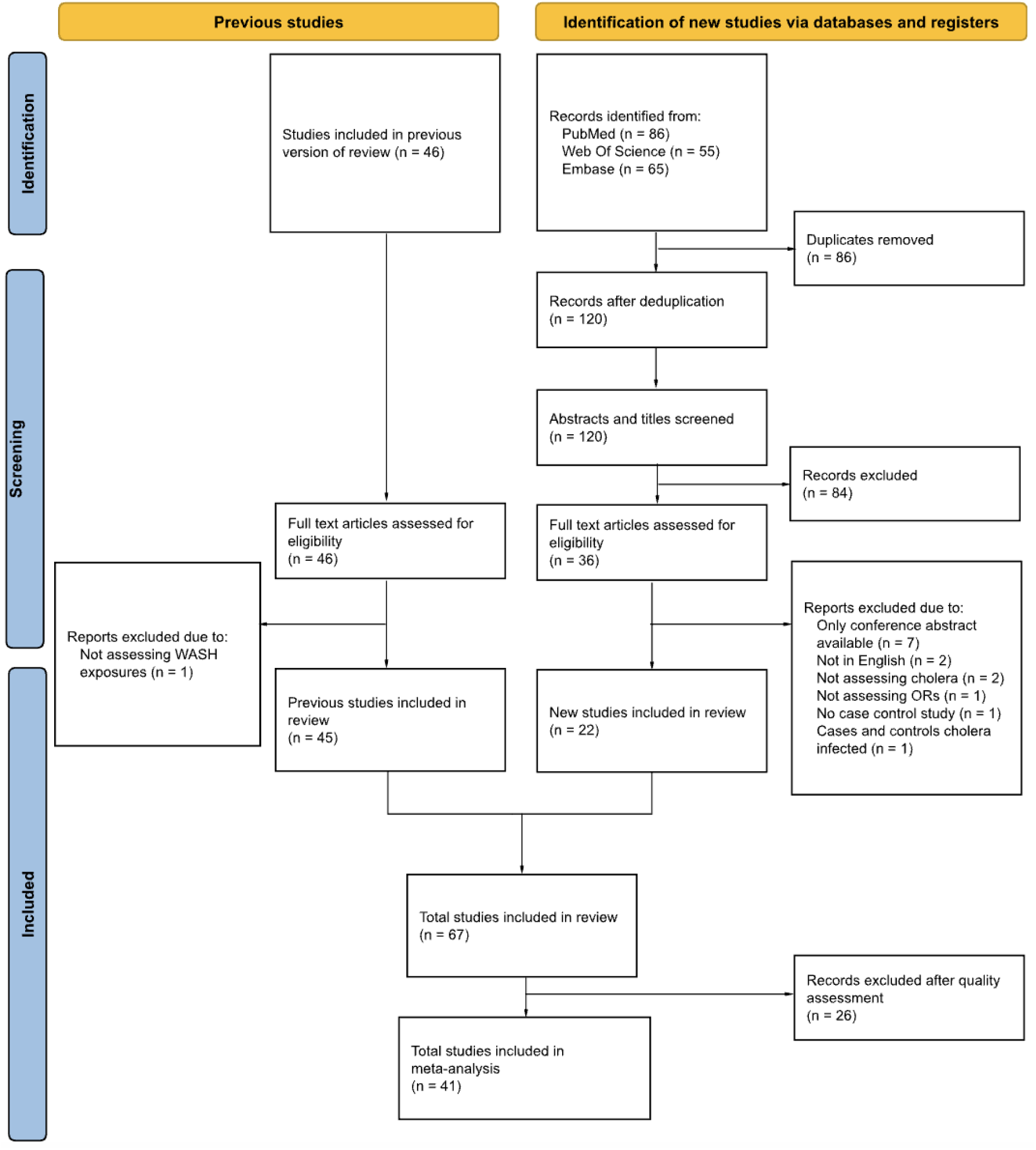
**PRISMA Flow Diagram.**

Wolfe *et al*. identified 46 studies in their review, which we also included [17]. During the evaluation of these articles, we noticed that one study focused on food exposures rather than WASH exposures. This article was therefore excluded.

Ultimately, 22 new studies in addition to 45 articles from the previous review of Wolfe *et al.* were included in this review. The total of 67 studies were then further assessed for quality.

### Characteristics of Included Studies

The newly identified studies were primarily conducted in Kenya, Uganda, and Ethiopia as well as in Yemen, Nigeria, Zambia, Vietnam, India, Ghana, and the Democratic Republic of the Congo [31–52]. The majority was conducted in Uganda with a total of six studies [37, 38, 43, 47, 48, 50], whereas the studies from the previous review were from 21 different countries with the majority of them conducted in Ethiopia, Haiti, India, Kenya, Malawi and Peru [53–97]. The combination of new and previously identified studies resulted in a wider geographical representation compared to the previous review. The diagnostic methods used to fully identify cholera cases were by culturing stool samples (39.7%), culturing rectal swabs (14.7%), collecting rectal swabs as well as stool cultures (14.7%), rapid test (7.4%), and rapid test as well as stool culture (2.9%). Only one study additionally collected blood samples and tested for antibody titres for cases as well as controls [97]. Two studies only collected blood samples for controls [84, 93] and one study only for a subset of controls [54]. Detailed descriptions of the characteristics of each study included can be found in S1 Table.

### Risk of Bias Assessment

Overall, the quality assessment with ROBINS-E resulted in 14 studies (20.9%) being classified as “some concerns”, 27 studies (40.3%) as “high risk”, and 26 studies (38.8%) as “very high risk” (Figs S1-2). The studies classified as “very high risk” were omitted from meta-analysis. Therefore, a total of 41 studies were included in the meta-analysis (Fig 1). Of the 45 previously included studies, 20 studies were excluded after our risk of bias assessment while the remaining 25 studies were included in the meta-analysis. Detailed description of the summarized results of the risk of assessment can be found in S1 Fig.

No publication bias was detected by inspecting the funnel plots (S4–S10 Figs) and Egger’s test (S2 Table).

### Results of Synthesis

A summary of the results of the meta-analysis can be found in Table 2.

**Table 2.** Results of the Meta-Analysis.

| Indicator of water quality | N exposure<br>s | Pooled OR<br>(95% CI) | Test of Heterogeneity |  |  | Wolfe et al.<br>(2018)<br>OR (95% CI) |
| --- | --- | --- | --- | --- | --- | --- |
| | | | Cochran's Q | $\tau^2$ | I<sup>2</sup><br>(95% CI) | |
| Improved water |  |  |  |  |  | 1.08<br>(0.54–2.15) |
| Safely managed water |  |  |  |  |  |  |
| Sachet water | 3 | 1.69<br>(1.13–2.52) | 3.505 | 0.000 | 0.00<br>(0–>99.56) |  |
| Bottled water | 5 | 0.52<br>(0.19–1.36) | 27.126** | 1.017 | 83.01<br>(53.62–97.80) | 0.35<br>(0.13–0.96) |
| Tap water | 11 | 0.92<br>(0.41–2.08) | 101.537** | 1.670 | 90.42<br>(79.65–96.84) |  |
| Basic water | 5 | 0.44<br>(0.27–0.69) | 7.460 | 0.008 | 2.49<br>(0.00–98.12) |  |
| Limited water | 4 | 0.84<br>(0.14–4.89) | 15.874** | 2.773 | 88.81<br>(58.13–99.33) |  |
| Unimproved water | 9 | 2.91<br>(1.21–7.02) | 52.609** | 1.437 | 88.67<br>(71.92–97.49) | 3.42<br>(2.47–4.74) |
| Surface Water | 10 | 3.40<br>(2.52–4.58) | 9.026 | 0.000 | 0.00<br>(0.00–75.97) | 2.27<br>(1.07–4.80)<sup>a</sup> |
| Treated water | 9 | 0.42<br>(0.27–0.65) | 19.214* | 0.231 | 54.21<br>(5.73–87.98) | 0.44<br>(0.35–0.56) |
| Boiled | 7 | 0.38<br>(0.17–0.84) | 24.006** | 0.899 | 81.52<br>(52.16–96.98) |  |
| Chlorinated | 6 | 0.37<br>(0.17–0.83) | 19.943** | 0.736 | 78.95<br>(40.66–96.66) |  |
| Observation of Treatment Materials | 4 | 0.41<br>(0.15–1.14) | 12.170** | 0.763 | 75.00<br>(19.80–98.15) |  |
| Untreated water | 15 | 2.51<br>(2.03–3.10) | 8.442 | 0.000 | 0.00<br>(0.00–34.40) | 3.47<br>(2.76–4.35) |
| Safe water storage | 6 | 0.19<br>(0.03 – 1.36) | 38.199** | 4.754 | 87.19<br>(63.92–97.62) | 0.55<br>(0.39–0.80) |
| Unsafe water storage | 5 | 1.32<br>(0.57 – 3.03) | 20.194** | 0.668 | 77.76<br>(37.99–97.08) | 2.79<br>(2.13–3.65) |

\*p < 0.05
\*\*p < 0.01
<sup>a</sup> categorized as surface water contact by Wolfe et al. [17]

### Water Source

A total of 21 exposures met the definition of safely managed water and were therefore further subdivided: sachet water, bottled water, and tap water. Sachet water consumption (n = 3) was associated with a 1.69 times higher risk in cholera (OR = 1.69, 95% CI: 1.13 to 2.52) (Fig 2). Bottled water (n = 5) was protective against cholera, but not statistically significant (OR = 0.52, 95% CI: 0.19 to 1.36). Similarly, tap water (n = 11) showed no significant protection against cholera (OR = 0.92, 95% CI: 0.41 to 2.08) (Fig 2).

**Fig 2.**
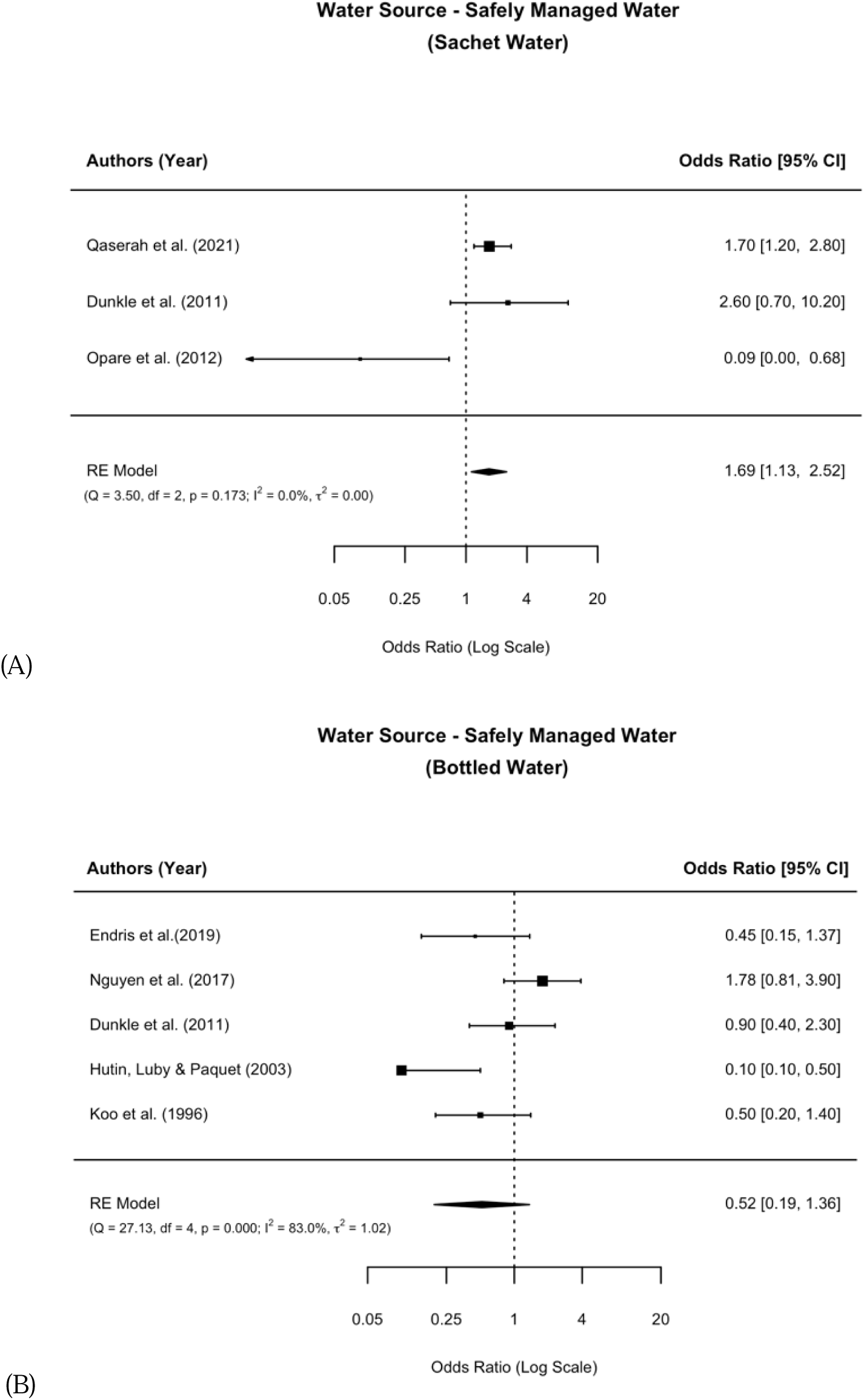
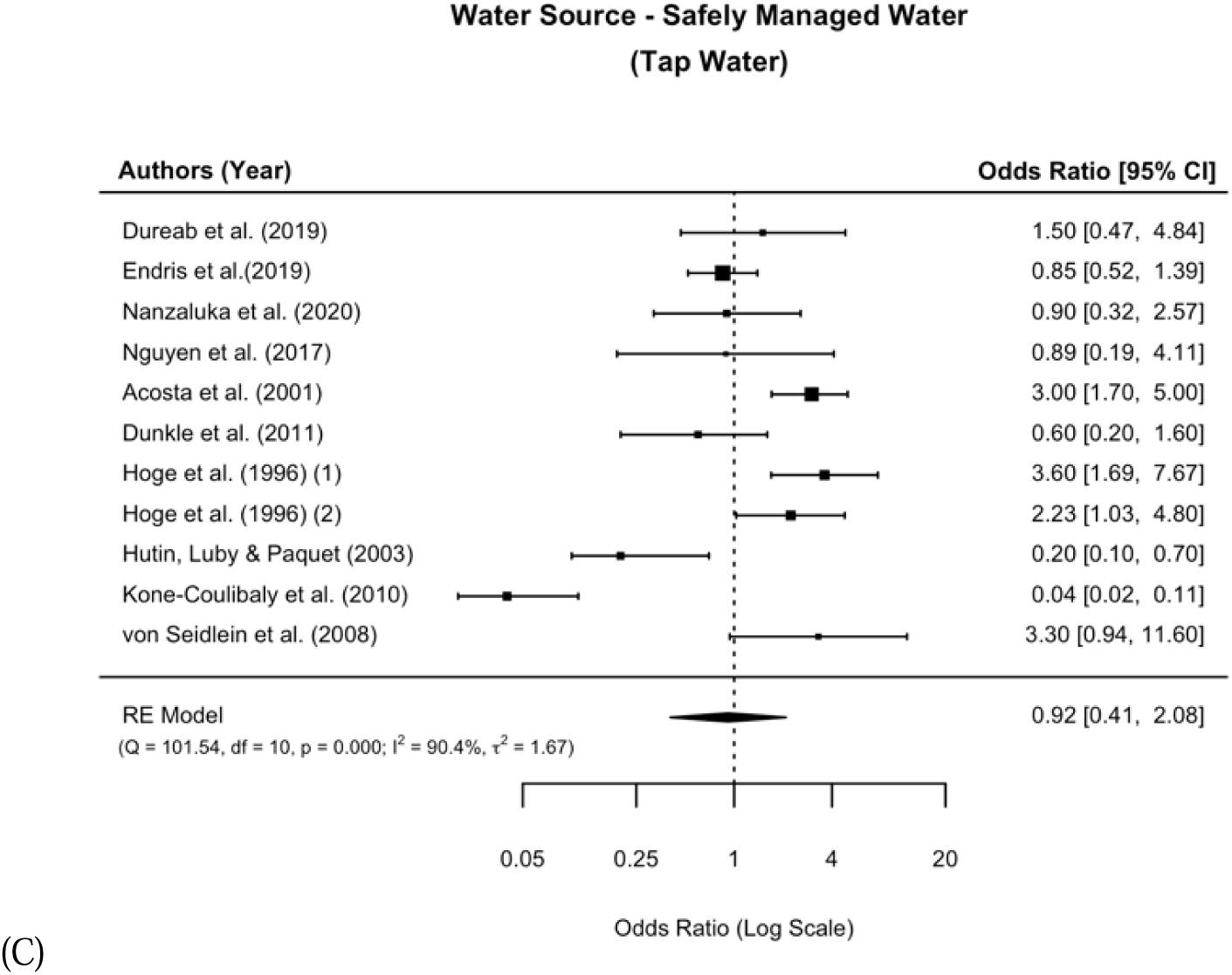
**Meta-analysis of the associations between the subgroups of safely managed water and cholera.**

For basic drinking water sources (n=5), a statistically significant protective effect against cholera was observed (OR=0.44, 95% CI: .27 to 0.69) (Fig 3).

**Fig 3.**
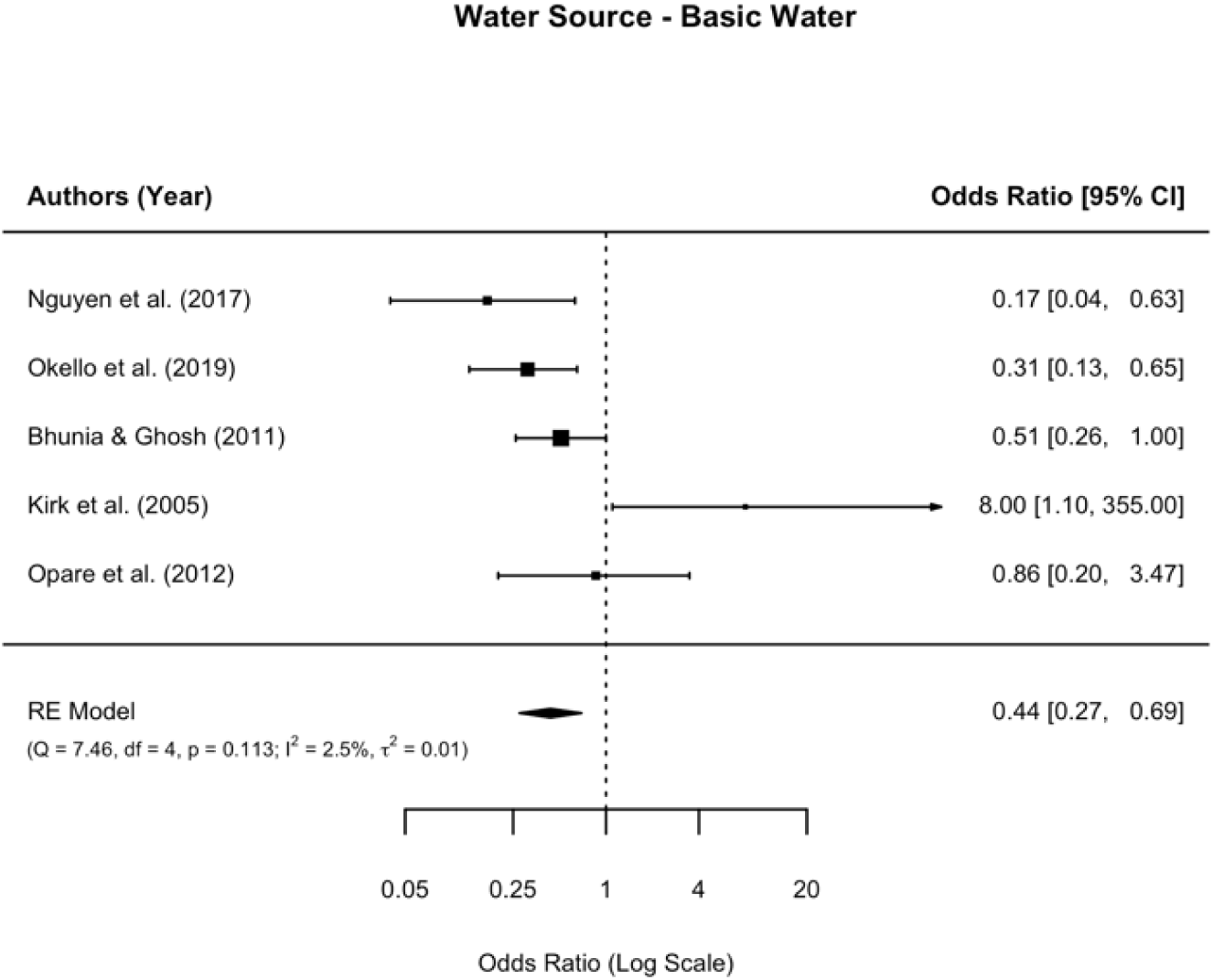
**Meta-analysis of the association between basic water and cholera.**

Limited water consumption did not show a significant association with cholera protection (n = 4, OR = 0.84, 95% CI: 0.14 to 4.89) (Fig S11).

However, the consumption of unimproved water, such as water from unprotected wells and surface water was associated with higher odds of cholera. Consuming unimproved drinking water (n = 9) resulted in increased odds of 2.91 (95% CI: 1.21 to 7.02) (Fig 4). Similarly, consuming surface water (n = 10) was significantly associated with cholera risk, with an OR of 3.40 (95% CI: 2.52 to 4.58), the highest odds for cholera compared to the other subgroups (Fig 4).

**Fig 4.**
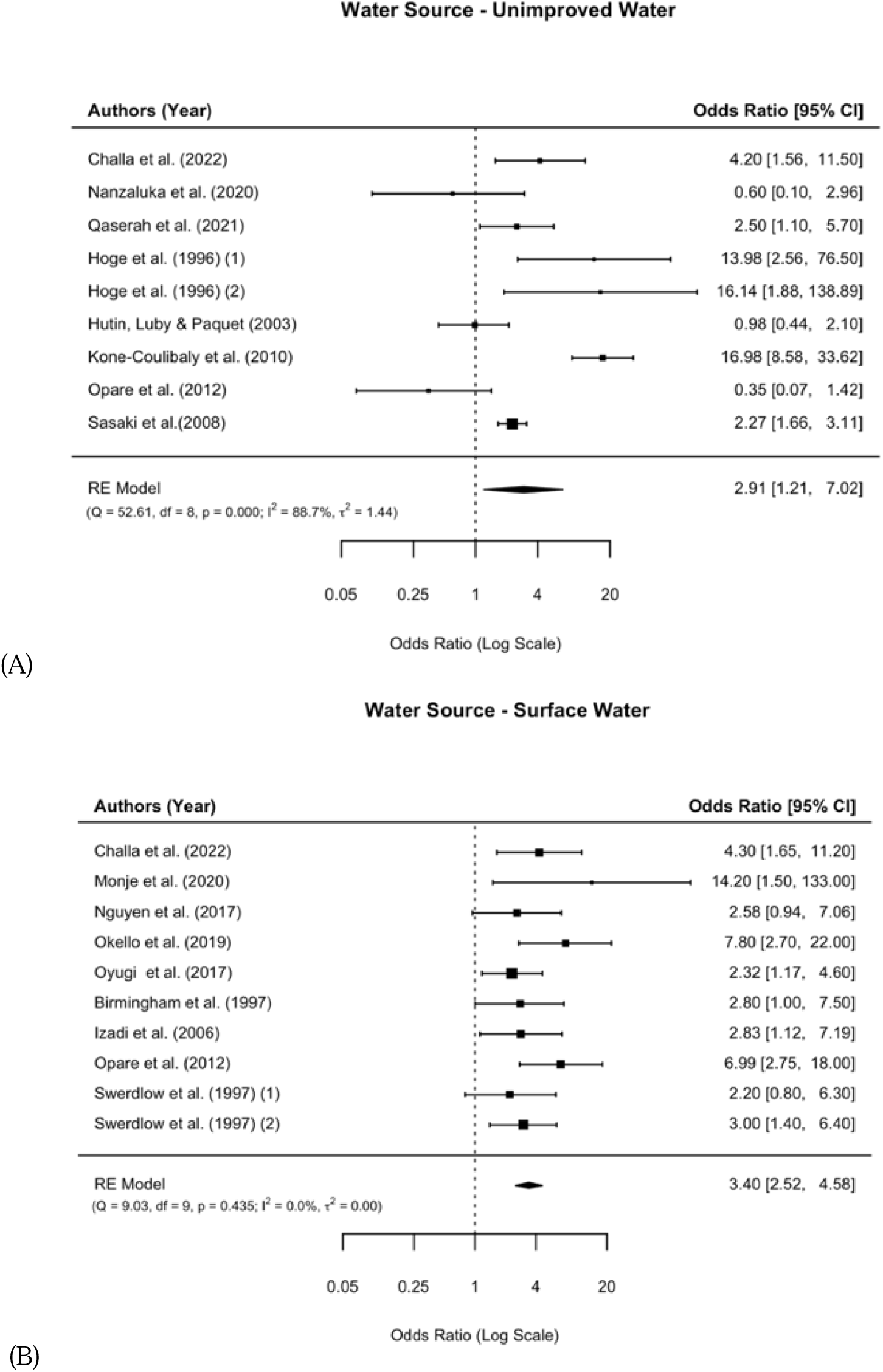
**Meta-analysis of the associations between unimproved water and cholera as well as surface water and cholera.**

### Water Treatment

Unspecified water treatment (n = 9) significantly decreased the odds of cholera by 58% (OR = 0.42, 95% CI: 0.27 to 0.65) (Fig S12). In contrast, untreated water (n = 15) significantly increased odds of cholera (OR = 2.51, 95% CI: 2.03 to 3.10) (Fig S12).

Similar results were obtained for the subgroups of treated water after synthesis. Boiling (n = 7) and chlorination of water (n = 6) significantly reduced odds for cholera with OR of 0.38 (95% CI: 0.17 to 0.84) and 0.37 (95% CI: 0.17 to 0.83), respectively (Fig 5). No significant protection was detected for having materials for water treatment observed (n = 4) (OR = 0.41, 95% CI: 0.15 to 1.14) (Fig 5).

**Fig 5.**
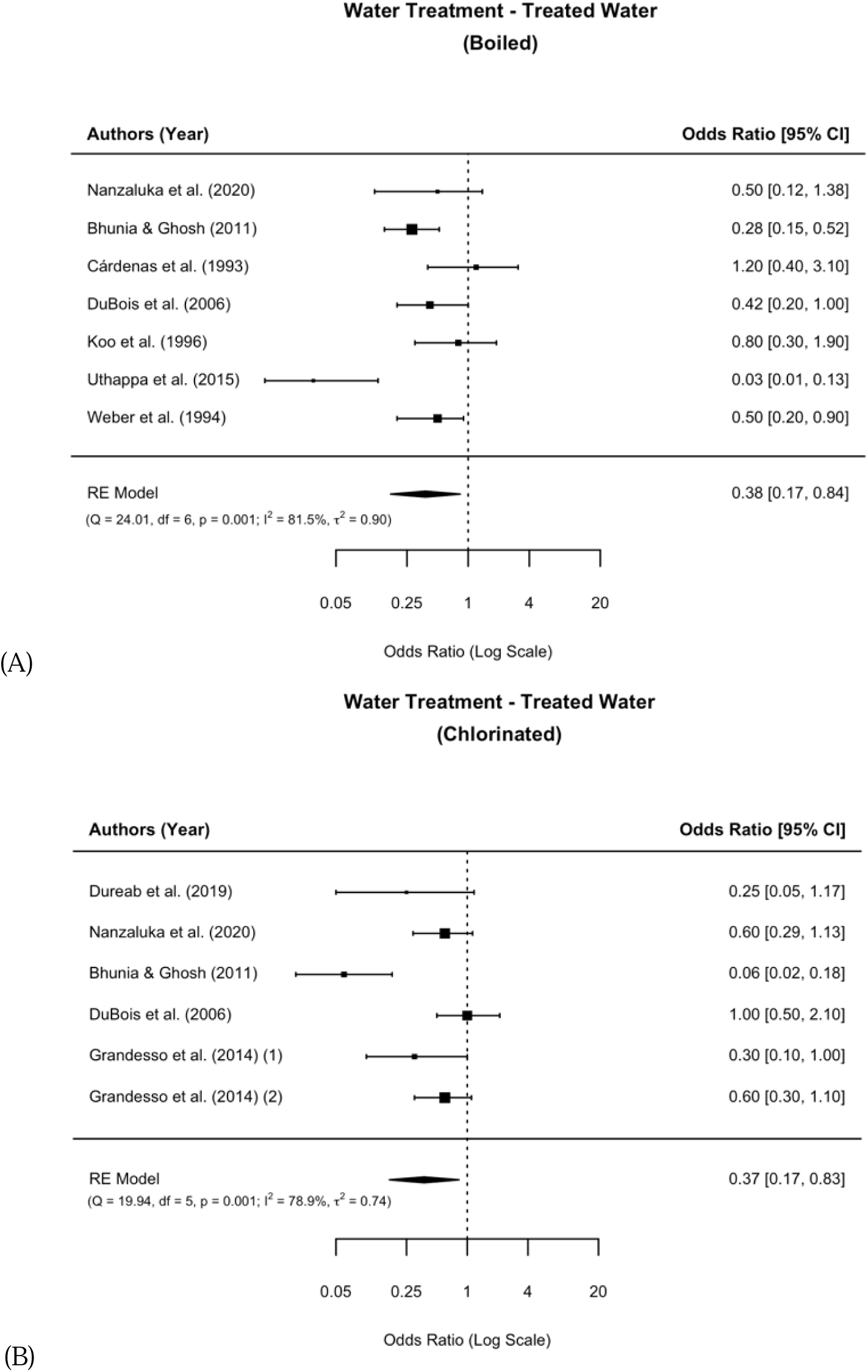
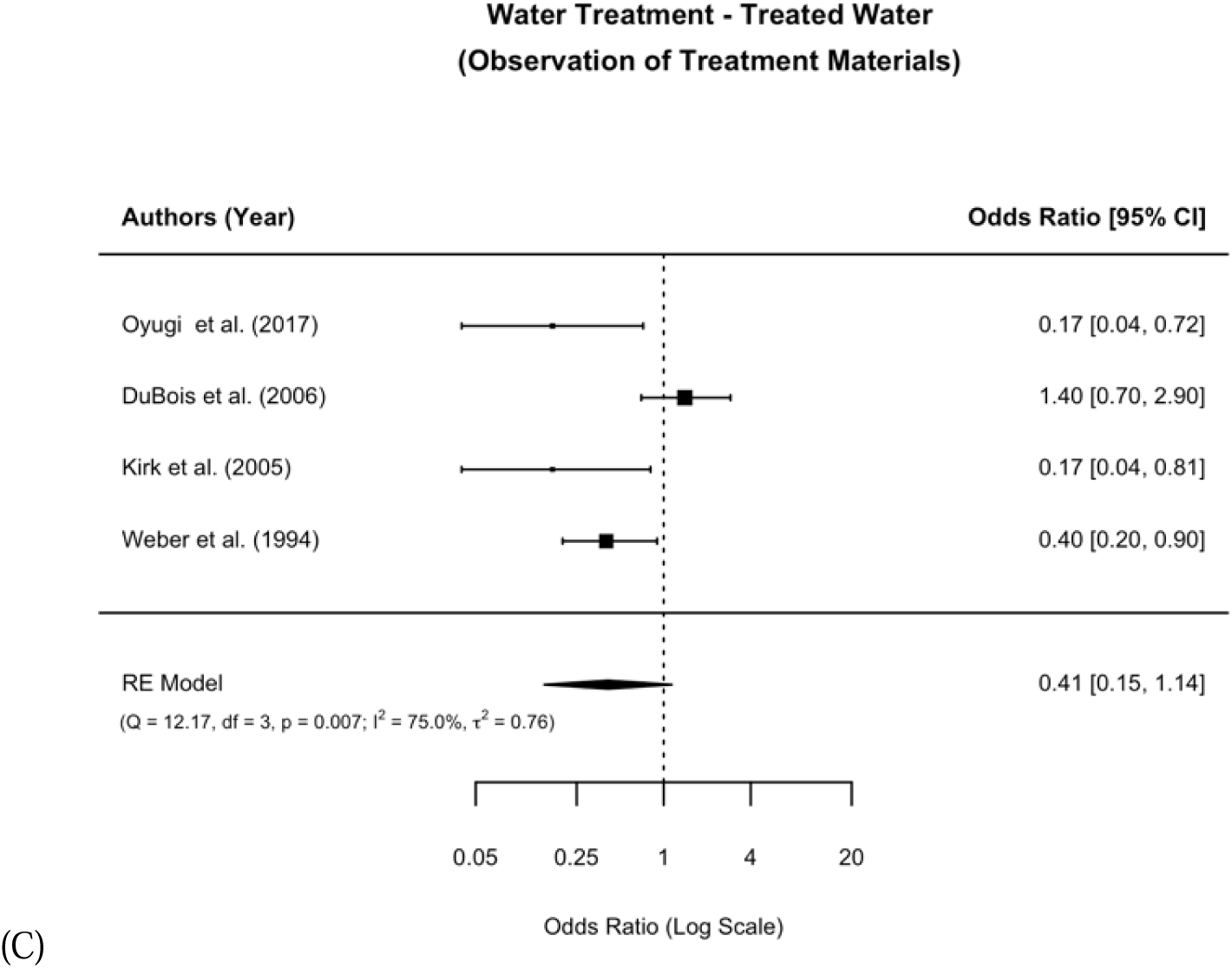
**Meta-analysis of the associations between water treatment and cholera.**

### Water Management

Safe water storage (n = 6) showed a non-significant reduction in OR of 0.19 (95% CI: 0.03 to 1.36) (Fig S13). Similarly, unsafe water storage (n = 5) had a non-significant increase in OR of 1.32 (95% CI: 0.57 to 3.03) (Fig S13).

### Heterogeneity

Substantial heterogeneity was observed for nearly all subgroups of water source other than consuming sachet water (I^2^ = 0.00%), basic water (I^2^ = 2.49%), and surface water (I^2^ = 0.00%) (Table 2). Similarly, only untreated water was found to have low heterogeneity in the water treatment category (I^2^ = 0.00%). Studies included on safe water storage as well as unsafe water storage revealed to be substantially heterogenous with I^2^ = 87.19% and I^2^ = 77.76%, respectively. Out of 14 conducted meta-analyses, heterogeneity was over I^2^ = 80% in 6 subgroups.

## Discussion

We conducted a systematic review and meta-analysis to examine the association between water quality and cholera. In particular, we updated the previous review by Wolfe *et al.* [17] by including data from case-control studies published from 2016 to 2022. Compared to the former review, a more rigorous risk of bias assessment was utilized as well as the formulation of more stringent inclusion criteria for meta-analysis. Our analyses revealed that most of the expected risk factors, such as the consumption of unimproved water and surface water, as well as the lack of water treatment, were associated with higher odds of cholera. Some of pooled estimates were substantially different from those in Wolfe *et al.* while qualitative insights were still the similar [17]. The exception was unsafe water storage, which failed to show statistical significance after being pooled. A similar lack of statistical significance, but for this case for a protective effect, was observed for safe storage of water. Water treatment and its subgroups showed significant protection against cholera, again in accordance with previous findings of Wolfe *et al.* [17]. However, the mere presence of materials for water treatment was not found to be statistically significant for protection. Contrary to expectations, the only subcategory of improved water that displayed significant protective associations against cholera was basic water. All other subcategories of improved water were not statistically significant. Although Wolfe *et al.* found a significant protective effect of drinking bottled water, our results could not replicate this observation [17]. Moreover, most striking was that the consumption of sachet water was unexpectedly associated with higher odds for cholera. Furthermore, large discrepancies in between-study heterogeneity was reported, as well as a wide ranges of observed ORs depending on the subcategory and subgroup.

Our findings confirm those of Wolfe *et al.* regarding the inconsistency of different subcategories of improved water [17]. It can be suggested that some supposedly safe water sources may not be protective but may in fact pose a higher risk than water sources that are lower on the JMP ladder, e.g., basic water or limited water, and may facilitate cholera outbreaks. We observed a number of studies in which municipal tap water was associated with higher odds of cholera [34, 53, 66, 96] as well as studies in which tap water was most likely contaminated [41, 57, 85, 91]. The reasons for this were often failures in the piping and sewage systems, which resulted in the tap water being contaminated by, for example, residues from open defecation [41, 52, 57]. In addition, water sold by vendors also appears to be at high risk of contamination. Contamination of packaged water sold on the street does also not appear uncommon, as evidenced by recent findings [41, 44]. Several studies have found significantly high levels of bacterial indicators in sachet water [98–100]. It appears that the level of contamination increases along the supply chain of the sachets [101]. There is evidence to suggests that the water sources used were not the underlying concern, but rather the packaging and handling of the sachets appeared to be strong indicators of contamination [102]. Given that some case-control studies failed to report protective effects of bottled water [36, 61, 72] or even showed increased odds [46], similar assumptions can presumably be made for bottled water. Packaged water in LMICs appear to often be of questionable quality, as modelling suggests [103]. However, the JMP categorization does not reflect this actual risk and the possibility of contamination (in its service ladder). This is especially concerning given the growing trend of consuming packaged water in LMICs as a result of water shortages and as an alternative to unimproved water sources [104, 105].

In contrast, the risk factors associated with cholera seem to be consistent and drive outbreaks. The consumption of surface as well as unimproved water tend to confer a constant risk of cholera. A similar pattern can be found in several other studies not exclusively focused on case-control studies [10, 106]. The homogeneity of exposures categorized as surface water consumption may support this. Consumption of surface water appears to be a risk factor regardless of setting and context of a study. Evidence of surface water being unsafe for drinking and increasing the risk for cholera and outbreaks can be found in past literature [107–109].

Furthermore, our results indicated that treating water with any method implies protection for cholera. Water treatment has the potential to prevent and slow down outbreaks across different LMICs, while eliminating the increased risk of cholera from consuming untreated water. This is in good agreement with Cohen and Colford who found a significant protection of boiling water against cholera and several other infectious diseases in similar setting [110]. The mere presence of equipment to treat water did not seem sufficient to provide significant protection. Appropriate training for correct and regular usage may be needed, as Lantagne and Yates previously described in their review, to fully take advantage of its protective effect [111].

The lack of a significant effect and the wide observed confidence interval for the either safe or unsafe storage of water could possibly stem from confounding factors such as where the water was collected from as suggested by Birmingham et al. in their case-control study [56]. Other important influencing factors can be whether the water was treated or the container itself was cleaned [14]. We observed that none of the exposures included for water management sufficiently controlled for these factors. Other reviews have in fact shown significance of an increased risk of unsafe water management and a lowered risk of safe water management [10, 106]. However, it seems that interventions focusing on the impact of safe water management have not been well-described yet and need further research [14].

There were several limitations to our review and meta-analysis that need be considered when interpreting our results. We only included studies that were peer-reviewed and published in English, excluding grey literature. Despite our efforts to reduce publication bias through statistical means, there may still be residual risk of publication bias.

In addition, a considerable degree of heterogeneity among the studies was observed in certain exposure subcategories and subgroups. Large differences in adjustment for confounders, measurement of exposure, methods of assessment of cases and controls, and contextual factors such as cultural differences, location, and time among the identified studies were most likely influential. It is unlikely that all these differences were adjusted for in the model selection of the meta-analysis. We often observed a lack of adjustment for confounders. Most studies did not sufficiently control for influencing effects of other WASH factors as well as other sources of drinking water. Also, the importance of a well-defined baseline risk of each exposure to assess ORs cannot be neglected in a study design since they vary across contexts and settings. However, a precise description was often missing [36, 46, 61, 72].

Furthermore, the decision to focus only on case-control studies as major evidence on the association between cholera and water can be found in case-control studies, it inherently results in a certain risk of recall bias due to its retrospective design.

Additionally, we observed that most studies either only culturally confirmed a subset of patients or identified cases solely by definition rather than testing. Therefore, when interpreting these results, a certain risk of misclassification must be considered given that many cholera cases are asymptomatic [112–114]. There is a possibility that the risk of bias assessment itself influenced the reported results. None of the included studies had a low risk of bias rating.

Another factor to consider when interpreting the results is that this review did not differentiate between endemic and epidemic cholera, which may influence the associations.

The quality assessment was more stringent in comparison to the previous review by Wolfe *et al.* due to the use of ROBINS-E, which was specifically designed for non-randomized studies with a particular focus on exposures [17, 23]. This allowed a more precise identification of studies that appeared to be of very high risk of bias. In addition, the quality of studies conducted since the previous review [17] did not noticeably improve. This resulted in the number of studies and exposures included in each subgroup and subcategory of the meta-analysis to be considerably smaller, which inevitably affects the power of this study.

In order to have enough studies to conduct the analysis, we used a modified definition of studies that were overall categorized as some concerns: Instead of just two domains, at least five domains had to be rated as some concern to be regarded as a high-risk study. Nevertheless, the number of studies that were classified as having some concerns and the corresponding exposures were not sufficient to allow a reasonable meta-analysis to be performed. This resulted in the inclusion of studies classified as high risk, which may has consequently biased our findings.

While these are important considerations to keep in mind when interpreting our results, we believe that our review provides a more profound understanding of the association between water quality and cholera. It lays the groundwork for future interventions as well as future research as cholera remains a major public health threat in LMICs. Simple and relatively inexpensive interventions such as water treatment have the potential to considerably reduce the burden of cholera infection. Regular water treatment can play a crucial role in prevention future outbreaks and should be considered as an early on containment measurement. This is particularly beneficial where water quality is compromised, and rapid intervention is needed. A holistic approach is essential for future interventions in LMICs: Safe water supply infrastructure must be expanded. It must be made accessible to people in LMICs who are most vulnerable to cholera, in order to avoid recourse to water sources with a high risk of infection. Moreover, it may not be sufficient to eliminate risk factors and rely on protective factors as containment measures for outbreaks. Water that is supposedly safely managed may be contaminated and, rather than being protective, may be a driver of outbreaks. Vended water is gaining importance in the face of water shortages [104] making it even more crucial to establish clear regulations to prevent contamination.

## Conclusion

It is critical to continue efforts to prevent and control cholera by improving the access to the safe water in addition to improving sanitation and hygiene. In addition, it would be necessary to make sure the so-called improved water sources such as sachet water or bottled water is safe in in LMICs. Relatively simple and inexpensive early protection measures such as boiling or chlorination must be considered to best prevent and contain future cholera outbreaks in LMICs.

## Data Availability

All data produced are available online at https://github.com/kimfinale/Cholera_WASH_meta.

## Acknowledgments

The authors appreciate Raphael Zellweger and Birkneh Tilahun Tedesse for their constructive comments on the final manuscript. We thank Nadja Kranz and Tim Paulus for their assistance in conceptional questions as well as their support in reviewing the manuscript. We would also like to thank Lisa Kaldich and Konstantinos Mechteridis for helping adjust the code when issues arose.

## Author Contributions

JHK conceptualised and designed the study. CK and TN initially searched the literature and assessed the included articles. GG and TN extracted all data. CK and TN analysed and interpreted the data. TN drafted the manuscript. JHK led the data extraction and analyses and interpretation of the results. All authors commented on the drafts of the manuscript and approved the final draft of the paper.

## Data Availability Statements

The dataset and R codes for generating figures and tables are available at the GitHub repository of the corresponding author: https://github.com/kimfinale/Cholera_WASH_meta

